# Views and experiences of Antimicrobial Stewardship interventions in paediatric secondary care settings: A Qualitative Evidence Synthesis

**DOI:** 10.1101/2024.05.29.24308153

**Authors:** Nirzer Mate, Stefania Vergnano, Christie Cabral

## Abstract

**Objectives:** Injudicious use of antimicrobial agents contributes to antimicrobial resistance. Antimicrobial Stewardship (AMS) interventions use strategies derived from evidence-based practices to ensure careful use of antibiotics. AMS is less common in paediatrics as compared to adult practice. As AMS success depends on organizational factors and individual behaviours, this study synthesizes the existing qualitative evidence exploring key barriers, facilitators, and acceptability of AMS.

**Methods:** *Design:* A systematic search of primary qualitative studies was conducted in electronic databases.

*Data sources:* MEDLINE, PsychINFO via OVID, CINAHL electronic database and handsearching of grey literature sources was done.

**Eligibility criteria:** Qualitative studies exploring parents” and/or clinicians” (doctors and nurses) views, attitudes, beliefs, and knowledge on antimicrobial stewardship programmes in paediatric and neonatal hospitals.

**Data extraction and synthesis:** Quality appraisal was done using the Critical Appraisal Skills Programme (CASP) tool for qualitative studies. The extracted data was then synthesised by drawing on meta-ethnography.

**Results:** A total of 6 studies met the inclusion criteria. 5 studies reported the views and experiences of doctors and nurses, and 1 study reported those of parents. The perceived value of AMS differed in neonatal and paediatric contexts. Structural barriers like resource allocation and hospital organization were a barrier to implementation and acceptability. Nurses reported a lack of formal education about AMS as a barrier.

**Conclusion/Implications:** The factors affecting AMS in paediatric secondary care vary with the stakeholders in question. This review identifies some of the factors that can be used to formulate service-level AMS interventions and programmes.

**Strengths and limitations of the study:**

- This is the first evidence synthesis of the qualitative literature exploring the beliefs and experiences of parents and clinicians regarding paediatric AMS.
- The facilitators and barriers were identified from themes representative of all the included studies, increasing their validity.
- While included studies were conducted across six countries, all were within the developed world which could limit the generalisability of the findings.
- A limited number of studies was included due to limited number of primary studies conducted in this area.

**Original protocol from PROSPERO:** Registration number: CRD42022346842

Available from: https://www.crd.york.ac.uk/prospero/display_record.php?ID=CRD42022346842 The process for data synthesis was changed from thematic analysis to meta-ethnography to accommodate the variety of ways in which the results were reported in the included studies.

## 1. Background

Antimicrobial resistance is a major public health concern, with a steady increase in emerging resistance worldwide.(1,2) Among others, injudicious use of antibiotics has been identified as one of the main culprits. To tackle this problem, antimicrobial stewardship (AMS) interventions are needed. AMS interventions are multidimensional strategies that target the known causes of injudicious use of antimicrobials, like over-prescription, non-compliance to clinical guidance and broad-spectrum antibiotic regimen, etc.(1,3) AMS can be achieved by using both persuasive (education, clinical guidelines, audit and feedback) and restrictive (formulary restriction, prior approval) strategies to change the prescribing behaviours of healthcare professionals.(3) The success of these interventions depends on organizational and behavioural factors (4)(5). It is, therefore, important to explore these factors to design and run a successful AMS programme.

Although most adult in-patients units in the UK have started implementing AMS programmes, paediatric programmes are falling behind. Important differences in the patient population have been identified, like the type of antimicrobial agents used and patient outcomes that are monitored (6) which influence the design, implementation and value of AMS interventions in paediatrics. The purpose of this study is to identify the key barriers and facilitators of implementing such interventions as well as to assess their acceptability in neonatal and paediatric hospitals.

## 2. Methods

Qualitative research aims to provide insight into the behaviours, views, experiences, and attitudes surrounding the study subject.(7) AMS programmes in paediatric settings are a new concept and therefore, not many studies have been conducted in this area. A qualitative evidence synthesis uses data from these primary qualitative studies to develop new knowledge. (8,9)

The review question was “What are the views and experiences of parents and clinicians regarding the implementation of antimicrobial stewardship programs in neonatal and paediatric secondary care settings?”

Due to the complexity of the research question, and the inclusion of a wider study population (clinicians and parent) meta-ethnography chosen as the synthesis design. Meta-ethnography allows for an in-depth and nuanced understanding of a phenomenon by bringing together the findings from multiple studies.(10) It can provide a comprehensive view of the research on a particular topic and can help to identify areas of agreement and disagreement among the studies.

### 2.1 Search Strategy and Inclusion criteria

Electronic databases were searched using free-text and MeSH terms relating to the study aims. Search terms for [antimicrobial stewardship] AND [paediatric or child or neonate] AND [parents or clinicians or nurses] AND [hospital or secondary care] AND [qualitative methods] were used. (Supplementary material 1) along with truncation and adjacency to optimize the sensitivity and specificity of the search. The electronic databases searched included Medline (OVID), CINAHL and PsychINFO and search terms were adapted to different databases. Additionally, handsearching was done in government websites, reference lists of included studies and inputs from relevant societies and interest groups.

Search limits were used to include papers from 2001 to 2022 (year the systematic review was conducted) This was done to accommodate the fact that the first global plans to address antibiotic resistance were published in 2001.(11)

Language was not limited at time of search.

The search results were imported to Rayyan software for systematic reviews (12)

The inclusion and exclusion criteria are tabulated in Table 1.

### 2.2 Data extraction and critical appraisal approach

Data extraction was conducted by NM and reviewed by CC and SV. Characteristics like aims of the study, study population, location, setting, methods of data collection, type of AMS activity and results were extracted and tabulated for organization and processing of the resultant data. (Table 2).

CASP (Critical appraisal skills programme) checklist (19) was used to assess the quality of the included studies (Table 3). All the selected studies fulfilled the requirements of the screening questions, and the quality was assessed and discussed.

### 2.3 Synthesis of findings/ Data Analysis

The synthesis used a modified Noblit and Hare’s meta ethnography process(20). Themes and quotes were identified from the results sections, within the scope of the research aim, keeping the supporting quotes with the subheadings where available, themes and quotes were extracted, organized it in tables and compared across the studies. Themes and quotes containing similar concepts were grouped together for a better understanding and cohesion. A summary description was written by coalescing similar or interdependent/related concepts. Contradictory data from different studies, but which related to a similar theme or concept was also grouped in these conceptual groups to maintain the richness of the data during synthesis.

This was an iterative process, with discussion between the researchers and ended when all the relevant themes identified in the studies were incorporated in the analysis.

## 3. RESULTS

The search resulted in 1186 hits, out of which 32 were eligible for full text screening. All of these were in English. Duplicate records were removed, the remaining studies underwent screening of their titles and abstracts by one researcher. In the next step, the resultant pool of studies underwent full-text screening that resulted in 6 final papers to be included in the study. (13–18)

All 6 met the inclusion criteria (Table 1,Figure 1) (21)

The main characteristics of the included studies are tabulated in Table 2. 5 studies were based in the USA(13,15–18) and 1 in Australia(14). Between the 6 studies, there were a total of 544 participants. While 3 studies explored the views of nurses(14,16,17), 2 focused on clinicians (medical director, senior attending, physicians, pharmacists)(13,15) and 1 explored the views of parents, children, and healthcare workers (18). However, this study had one section on antimicrobial stewardship and only reported parents” input on it. 2 studies were set in Neonatal Units (13,15), 3 were set in paediatric hospitals(14,16,17) and 1 was set in a paediatric oncology department. (18)

Quality appraisal of the studies is presented in Table 3.

### Synthesis

Nine themes emerged from the included studies.

Five themes were seen commonly in the included studies, namely, Value of AMS, Structural barriers, Multidisciplinary collaboration and Communication, Nurses’ role in AMS, and Knowledge gap.

The remaining four themes were: Perceived low and narrow-spectrum consumption, Parental influence, Champion representation, and Monitoring and feedback. These are summarized in Table 4, along with the supporting statements from the results of the included studies.

#### 1. Value of AMS

In neonatal units (NNUs), the participants had contrasting opinions regarding the value of AMS. Physician participants in one study (15) viewed the collaboration of AMS team in rounds as disruptive, stating that the “presence of a pharmacist was enough”. On the contrary, Antimicrobial Stewardship Collaboration (ASC) was seen as “valuable and important in reducing antibiotic usage rates and practice variation.” by participants in another study(13).

> “We tried it at one point but we couldn”t get the neos [neonatologists] to round with us. The feedback we got was that it was more disruptive than helpful.” [Physicians] (15)

In paediatric hospitals, the value of AMS was reported as contextual. Healthcare professionals stated that the choice of antibiotics and the choice of population influenced the value of antimicrobial resistance in different paediatric units(18). For example, antimicrobial resistance did not have an impact on individual decision making about infection prophylaxis. (18) Additionally, acutely sick status of patients in secondary care, and difficulty in changing the route of administration (especially from intravenous to oral, due to palatability and/or adherence) made it difficult to implement AMS in paediatric wards.(17)

#### 2. Structural barriers

The second category, structural barriers, addressed the barriers at the organizational level, out of the scope of the clinicians. This includes resource allocation, time, human resources, finances, and technological resources (13,15,16). Institutional structure, like the affiliation of the NNU to a parent organization, influenced the AMS coverage. For example, some NNUs were part of a general hospital, instead of a children’s hospital and therefore, did not receive AMS guidance from either of the two.(15) Resource allocation, especially time was a barrier in getting engagement and in implementing unit change.(13)(15)

#### 3. Multidisciplinary collaboration and Communication

This category talks about the various barriers and drivers in the context of multidisciplinary teams and collaboration with different stakeholders. Neonatologists were resistant to input from the AMS team. (15)The recommendations came from adult AMS providers who lacked paediatric expertise. One study reported that the nursing team preferred to follow the doctors and physicians” guidance over that of the AMS team(15). They felt reluctant to defer, question and provide input to treatment plans due to reasons such as prescriber pushback, lack of involvement by consulting services, and duplication of work between the clinical team members(15–17). Unit culture and multidisciplinary collaboration were drivers in facilitating change(13). Relations between the external facilitators and the NICU affected the effectiveness of the AMS programme, with supporting the familiarity among the team members as a driver towards successful collaboration(13).

Communication barriers were reported by different participant populations in their own settings. Lack of communication between the nursing and AMS teams, in one case, resulted in unawareness about existing AMS hospital policies altogether(14). Collaboration with pharmacists to support daily rounds or creating antibiotic timeouts was seen as useful(13).

> “I’ve never been called by our nursery. It’s not a priority for us.” [AMS programme provider](15)

#### 4. Nurses” involvement in AMS

The role of nurses in AMS was seen as valuable (14,16,17), but lacking recognition.(14) Some strategies suggested by the AMS team were deemed outside the scope of nurses’ practice by the participants with the risks of duplication of work and resultant waste of resources. (17) Nurses believed their role was to advocate for the patients if they were uncomfortable with the physician’s treatment plans (14,16), however, some participants felt that they weren’t very good at questioning these plans.(14) Patient advocacy was exercised by providing individualized care, facilitating a positive patient experience, ensuring the best route of administration, and ensuring timelines. Advocacy extended to the carers/parents of children. (14)

Paediatric nurse participants believed that they had an important role to play in educating caregivers about the AMS strategies, the rationale around them, and providing tailored guidance. (14,16)

> “We [are] not very good, I don’t think we are very good at questioning why a child is on antibiotics. We’ll often look at a medical chart and go, “Oh, why are they on this?’or’Oh, they must just want to keep it going.’ You know or they might just want to cover with it, so sometimes I think we overdo it, but as nurses we don’t question antibiotic use as much as we probably should do” [Nurse] (14)

#### 5. Education

The presence of a “knowledge gap’ was mentioned several times in this group of studies, with participants expressing the need for formal training about AMS, including recommended techniques, AMS terminology, management strategies and antibiotic dosages and strength. (14,16,17). Paediatric nurses desired formal education about AMS and reported filling knowledge gaps through informal learning like reading drug formularies, reviewing latest evidence-based practices and having discussions on rounds. (16) Participants expressed a desire to have a community of shared learning within the collaborative. (13)

> “If I have never heard of it, I looked it up and I learn. That’s how I learned most of my antibiotic knowledge.”[Nurse] (16)

### 6. Other

#### 6.1 Perceived low consumption & Narrow spectrum consumption

This theme was identified from a study in the NICU setting, where AMS providers cited specific patient population and care practices as barriers to AMS implementation as the programme focused on adult patients.(15) The neonatal units did not use broad spectrum antibiotics, which was a focus of the AMS (15).

#### 6.2 Parental influence

Nurses in paediatric wards faced some pushback from parents when implementing certain strategies, like encouraging non-invasive methods to obtain culture samples where invasive methods were indicated(18).While they did acknowledge the possibility of development of resistance, parents and healthcare workers in paediatric oncology department viewed antimicrobial resistance as an issue on the community level, rather than the individual level. (18)

#### 6.3 Champion representation

This theme was reported in both the studies set in NNUs. The involvement of a champion representative from the clinical team reduced resistance to AMS recommendations and increased buy-in from clinicians(15) and Antimicrobial Stewardship Collaborations witnessed high levels of engagement from physician and nurse champion (13). However, the engagement was limited to the level of local leadership, not reaching the individual clinician level. It was noteworthy that this individual capacity to change influenced the facilitation of AMS, making it a potential target for behaviour change interventions. (13)

#### 6.4 Feedback and monitoring

Monitoring, evaluation, and feedback was perceived as important in NNU settings(13). Lack of accountability to follow guidelines about proper techniques in some AMS interventions in paediatric wards was seen as a barrier to implementation(17).

## 4. DISCUSSION

This qualitative evidence synthesis found that the perceived value of AMS was one of the main factors that affected the acceptability of AMS in this setting. Where AMS value was recognised there was good understanding and adherence otherwise it was seen as an unhelpful intrusion. Structural organization, multidisciplinary collaboration and communication, and nurses’ involvement in AMS are the prominent opportunity that could drive behaviour change around AMS in paediatrics.

A consistent need for structural reform has emerged from previous research, which showed that most children’s hospitals in UK, delivered AMS with limited formal funding. (23) Structural reforms in terms of technological resources, (like electronic systems to monitor antibiotic use), updating drug charts, and manpower (6), all require financial support. Recognition of AMS in paediatrics is essential to secure consistent funding.

Patient care and treatment is coordinated by a multidisciplinary, interprofessional team, with effective communication playing an important role in smooth execution of AMS strategies (24). Consequently, leadership and hierarchy within the team has been found to influence prescribing patterns in wards. (26) Research on the social factors surrounding the implementation of AMS also suggest that communication, and relationship between stakeholders has a significant impact on AMS buy-in. (26).

The results indicate that pressure from parents/carers to undertake or omit certain AMS activities was an important extrinsic factor affecting the implementation of AMS. This influence comes from a confidence in antibiotic therapy, fear of more serious complications or perceived discomfort to the child (17,27). Prescribers are less likely to continue antibiotics in the absence of external pressure (28), further supporting the usefulness of educating nurses and caregivers about antimicrobial resistance and AMS and promoting input and buy-in from these groups when designing AMS programmes.

In the neonatal population it was felt that general AMS rules did not necessarily apply and that there would be need to adapt the AMS strategies to the needs of this special population, it was felt that neonatal specialists would need to be involved in the AMS decisions.

This review identifies factors that are unique to paediatric and neonatal units. It highlights the possibility and express the need for primary research to look at strategies that strengthen the links between antimicrobial stewardship and clinical paediatric and neonatal teams. By incorporating input from a diverse population, the results can act as a well-rounded set of frameworks for building patient safety and public health policies in paediatric hospitals and neonatal units.

This study draws on the experience of multiple stakeholders in secondary paediatric care, including AMS service providers, physicians, nurses, and parents. The search was designed to result in a wide variety of participant populations in various contexts, enabling the comparison of collected evidence in a comprehensive manner.

The use of meta-ethnography in synthesis of findings allows for the translation of one study into another, overcoming the difference in the ways the results were presented within the group of studies. (20) The systematic review methodology provides a unique advantage of identifying gaps in research and areas where further research is needed. This can be especially helpful while designing further studies in this field.

The qualitative studies (n=6) were limited, and some were not of very good quality.

The search strategy may have missed studies that used terms other than those included in the database searches, however this was reviewed by experienced clinicians and a librarian. Most of the included studies were from the USA and one from Australia, which could limit the acceptability of the findings to other settings.

## Conclusions

This QES shows that the attitudes of clinicians towards paediatric AMS was contextual and varied with the AMS recommendations. This suggests that AMS interventions need to be tailored according to the setting they are intended for, with input from all stakeholders in the setting. (6)Multidisciplinary collaboration came out to be central to implementation and acceptability of the interventions. This was reiterated by a call for effective communication within and outside the clinical team. Some unique structural barriers (institutional affiliation) were discovered in the context of paediatric care among others that were common to other departments, such as time and human resource allocation. Parental pressure, lack of knowledge and individual capacity to change emerged as common barriers to implementation of AMS strategies at the patient level.

This QES can act as a useful starting point for formulating AMS strategies in Quality Improvement and Patient Safety. It has produced theories in the form of hypotheses that could be tested by other researchers. This study highlights the difference in value and application of AMS in neonates and paediatric populations, indicating the need of separate research in these populations.

## Supporting information

PRISMA flowchart

## Data Availability

All data produced in the present work are contained in the manuscript.

## Acknowledgements

I would like to thank the University of Bristol for giving me the opportunity to find my interest in qualitative research through the Masters in Public Health degree programme. I would also like to thank friends, family and colleagues for their support and well wishes.

## REFERENCES

1. Antimicrobial resistance [Internet]. [cited 2022 Aug 31]. Available from: https://www.who.int/news-room/fact-sheets/detail/antimicrobial-resistance

2. So A, Furlong M, Heddini A. Globalisation and antibiotic resistance. BMJ [Internet]. 2010 Sep 21 [cited 2022 Aug 31];341(7774):615. Available from: https://www.bmj.com/content/341/bmj.c5116

3. Fishman N. Antimicrobial stewardship. Am J Infect Control. 2006 Jun;34(5 SUPPL.).

4. van der Geest S, Whyte SR, Hardon A. THE ANTHROPOLOGY OF PHARMACEUTICALS: A Biographical Approach. 10.1146/annurev.anthro251153 [Internet]. 2003 Nov 28 [cited 2022 Aug 31];25(1):153–78. Available from: https://www.annualreviews.org/doi/abs/10.1146/annurev.anthro.25.1.153

5. Charani E, Castro-Sánchez E, Holmes A. The role of behavior change in antimicrobial stewardship. Vol. 28, Infectious Disease Clinics of North America. W.B. Saunders; 2014. p. 169–75.

6. Newland JG, Hersh AL. Purpose and design of antimicrobial stewardship programs in pediatrics. Pediatric Infectious Disease Journal [Internet]. 2010 [cited 2022 Aug 31];29(9):862–3. Available from: https://journals.lww.com/pidj/Fulltext/2010/09000/Purpose_and_Design_of_Antimicrobial_Stewardship.18.aspx

7. Tong A, Flemming K, McInnes E, Oliver S, Craig J. Enhancing transparency in reporting the synthesis of qualitative research: ENTREQ. BMC Med Res Methodol [Internet]. 2012 [cited 2022 Aug 26];12. Available from: https://pubmed.ncbi.nlm.nih.gov/23185978/

8. Britten N, Campbell R, Pope C, Donovan J, Morgan M, Pill R. Using meta ethnography to synthesise qualitative research: a worked example. J Health Serv Res Policy [Internet]. 2002 Oct [cited 2022 Aug 31];7(4):209–15. Available from: https://pubmed.ncbi.nlm.nih.gov/12425780/

9. Flemming K, Noyes J. Qualitative Evidence Synthesis: Where Are We at? Int J Qual Methods. 2021;20.

10. Britten N, Campbell R, Pope C, Donovan J, Morgan M, Pill R. Using meta ethnography to synthesise qualitative research: a worked example. Vol. 7, Journal of Health Services Research & Policy. 2002.

11. WHO Global Strategy for Containment of Antimicrobial Resistance. 2001;

12. Ouzzani M, Hammady H, Fedorowicz Z, Elmagarmid A. Rayyan-a web and mobile app for systematic reviews. Syst Rev [Internet]. 2016 Dec 5 [cited 2022 Sep 2];5(1). Available from: https://www.rayyan.ai/cite/

13. Quinn JM, Gephart SM, Davis MP. External Facilitation as an Evidence-Based Practice Implementation Strategy During an Antibiotic Stewardship Collaborative in Neonatal Intensive Care Units. Vol. 16, Worldviews on Evidence-Based Nursing. 2000.

14. Kilpatrick M, Bouchoucha SL, Hutchinson A. Antimicrobial stewardship and infection prevention and control in atopic dermatitis in children. Am J Infect Control. 2019 Jun 1;47(6):720–2.

15. Cantey JB, Vora N, Sunkara M. Prevalence, Characteristics, and Perception of Nursery Antibiotic Stewardship Coverage in the United States. J Pediatric Infect Dis Soc [Internet]. 2016 Jul 15;6(3):piw040. Available from: https://academic.oup.com/jpids/article-lookup/doi/10.1093/jpids/piw040

16. Hamdy RF, Neal W, Nicholson L, Ansusinha E, King S. Pediatric nurses’ perceptions of their role in antimicrobial stewardship: A focus group study. J Pediatr Nurs. 2019 Sep 1;48:10–7.

17. Carter EJ, Greendyke WG, Furuya EY, Srinivasan A, Shelley AN, Bothra A, et al. Exploring the nurses’ role in antibiotic stewardship: A multisite qualitative study of nurses and infection preventionists. Am J Infect Control. 2018 May 1;46(5):492–7.

18. Diorio C, Tomlinson D, Boydell KM, Regier DA, Ethier MC, Alli A, et al. Attitudes toward Infection Prophylaxis in Pediatric Oncology: A Qualitative Approach. PLoS One. 2012 Oct 24;7(10).

19. Critical Appraisal Skills Programme. CASP Qualitative Checklist. [cited 2022 Aug 26]; Available from: https://casp-uk.net/casp-tools-checklists/

20. Noblit GW, Hare RDwight. Meta-ethnography□: synthesizing qualitative studies. 1988;88.

21. Page MJ, McKenzie JE, Bossuyt PM, Boutron I, Hoffmann TC, Mulrow CD, et al. The PRISMA 2020 statement: An updated guideline for reporting systematic reviews. The BMJ. 2021 Mar 29;372.

22. Patel SJ, Saiman L. Principles and Strategies of Antimicrobial Stewardship in the Neonatal Intensive Care Unit. Semin Perinatol. 2012 Dec;36(6):431–6.

23. Vergnano S, Bamford A, Bandi S, Chappel F, Demirjian A, Doerholt K, et al. Paediatric antimicrobial stewardship programmes in the UK’s regional children’s hospitals. Journal of Hospital Infection [Internet]. 2020 Aug 1 [cited 2022 Sep 11];105(4):736–40. Available from: http://www.journalofhospitalinfection.com/article/S0195670120302619/fulltext

24. Principi N, Esposito S. Antimicrobial stewardship in paediatrics. BMC Infect Dis [Internet]. 2016 Aug 18 [cited 2022 Aug 31];16(1):1–8. Available from: https://link.springer.com/articles/10.1186/s12879-016-1772-z

25. Lewis PJ, Tully MP. Uncomfortable prescribing decisions in hospitals: the impact of teamwork. J R Soc Med [Internet]. 2009 Nov 11 [cited 2022 Aug 31];102(11):481. Available from: /pmc/articles/PMC2770359/

26. Pakyz AL, Moczygemba LR, Vanderwielen LM, Edmond MB, Stevens MP, Kuzel AJ. Facilitators and barriers to implementing antimicrobial stewardship strategies: Results from a qualitative study. Am J Infect Control. 2014 Oct 1;42(10):S257–63.

27. Teixeira Rodrigues A, Roque F, Falcão A, Figueiras A, Herdeiro MT. Understanding physician antibiotic prescribing behaviour: a systematic review of qualitative studies. Int J Antimicrob Agents. 2013 Mar 1;41(3):203–12.

28. Roope LSJ, Buchanan J, Morrell L, Pouwels KB, Sivyer K, Mowbray F, et al. Why do hospital prescribers continue antibiotics when it is safe to stop? Results of a choice experiment survey. BMC Med [Internet]. 2020 Jul 30 [cited 2022 Sep 11];18(1):1–11. Available from: https://bmcmedicine.biomedcentral.com/articles/10.1186/s12916-020-01660-4

