## Supplementary material for "Views and experiences of Antimicrobial Stewardship interventions in paediatric secondary care settings: A Qualitative Evidence Synthesis"

**TABLES**

Table 1: Study inclusion and exclusion criteria:

| **Characteristic** | **Inclusion criteria** | **Exclusion criteria** |
| --- | --- | --- |
| **Study focus** | Explored parents’ and clinicians’ (doctors and nurses) views, attitudes, beliefs, and knowledge on antimicrobial stewardship programmes in paediatric and neonatal hospitals (secondary care). |  |
| **Design** | Used qualitative methods (e.g. interviews, focus groups, observation, etc) to collect and analyse data. Studies had to report themes or summary of finding, with or without quotes. | Included only quantitative data (quantitative analysis of survey response) or collected data using questionnaires. |
| **Setting** | Reported data collected from paediatric secondary care (Neonatal units, Paediatric ICUs, General Paediatric wards, etc) in any country | Reported data from primary care (General or Family practices) |
| **Population** | a) Defined paediatric population as those between zero to 18 years of age.  (b) Included parents and clinicians with experience with antimicrobial stewardship activities and programs in paediatric settings.  (c) Included both adults and paediatric age group and reported findings in relation to the paediatric or neonatal group separately. |  |

Table 2: Description of included studies (grouped by study population)

| **First author, Date** | **Aim** | **Country** | **Setting** | **Data collection** | **Population** | **Type of AMS Intervention** |
| --- | --- | --- | --- | --- | --- | --- |
| Cantey,2017 (16) | To describe the characteristics of nurseries with and without ASP coverage and to determine perceptions of and barriers to nursery ASP coverage. | USA | NICU | Semi-structured interviews | 1. Nursery provider: medical director, senior attending  2. ASP provider: Pharmacist, physician | One or more infectious diseases physician or clinical pharmacist with infectious diseases training who  1) Oversee antibiotic prescribing and provide feedback within their centre 2) Receive some fraction of FTE compensation for their stewardship activities |
| Carter, 2018 (18) | Exploring the nurses’ role in antibiotic stewardship: A multisite qualitative study of nurses and infection preventionist | USA | Paediatric Hospital | Focus Groups,  Semi-structured interviews | Nurses (adult and paediatric),  Nurse managers,  Infection preventionists | 1) Questioning the need for urine cultures;  2) Ensuring proper culturing technique;  3) Recording an accurate penicillin drug allergy history;  4) Encouraging the prompt transition from intravenous (IV) to oral (PO) antibiotics 5) Initiating an antibiotic timeout. |
| Hamdy, 2019(17) | To explore paediatric nurses' perceptions of their role in antimicrobial stewardship | USA | Children’s Hospital | Focus Groups | Paediatric Nurses | 1) Developing and implementing institution-specific clinical pathways detailing choice of antibiotics for specific conditions  2) Identifying opportunities to optimize drug, dose, duration, and route of antimicrobial therapy  3) Providing education to providers, patients, and their families on appropriate antibiotic use  4) Restricting prescribing for selected antimicrobials |
| Kilpatrick, 2019 (15) | To explore the perceptions of dermatology nurses on their role in antimicrobial stewardship when caring for children with atopic dermatitis. | Australia | Tertiary Metropolitan Children's Hospital: Emergency dept,  short stay medical ward and dermatology department | Semi-structured interviews, Focus groups | Paediatric Nurses | AMS in Atopic Dermatitis |
| Quinn, 2019(14) | To categorize and describe essential features of facilitation in the context of implementing an EBP using perspectives elicited from neonatal clinicians and external facilitators (EFs). | USA | NICU | Semi-structured interviews | Neonatal clinicians  External Facilitators | External Facilitation as AMS implementation strategy |
| Diorio, 2012 (19) | To describe the attitudes of key stakeholders (parents, children, healthcare professionals) towards infection prophylaxis in paediatric oncology | USA | Paediatric Oncology dept | Semi-structured interviews | Parents  Children  Healthcare workers | No AMS activity |

Table 3: Quality Appraisal using Critical Appraisal Skills Programme (CASP)(19) tool

| **Sr no** | **CASP tool questions** | **First Author** | | | | | | |
| --- | --- | --- | --- | --- | --- | --- | --- | --- |
|  |  | *Cantey*(16) | *Carter*(18) | *Hamdy*(17) | | *Kilpatrick*(15) | *Quinn*(14) | *Diorio*(19) |
| 1 | Was there a clear statement of the aims of the research? | Yes | Yes | Yes | Yes | | Yes | Yes |
| 2 | Is a qualitative methodology appropriate? | Yes | Yes | Yes | Yes | | Yes | Yes |
| 3 | Was the research design appropriate to address the aims of the research? | No | Yes | Yes | Yes | | Yes | Yes |
| 4 | Was the recruitment strategy appropriate to the aims of the research? | Yes | Yes | Yes | No | | Yes | Yes |
| 5 | Was the data collected in a way that addressed the research issue? | Can't tell | Yes | Yes | Yes | | Yes | Yes |
| 6 | Has the relationship between researcher and participants been adequately considered? | No | No | No | No | | No | No |
| 7 | Have ethical issues been taken into consideration? | Can't tell | Yes | Yes | Yes | | Yes | Yes |
| 8 | Was the data analysis sufficiently rigorous? | Yes | Yes | Yes | No | | Yes | Yes |
| 9 | Is there a clear statement of findings? | Yes | Yes | Yes | No | | Yes | No |
| 10 | How valuable is the research? | Yes | Yes | Yes | No | | Yes | Yes |

Table 4: Summary of synthesis

| **Summary of synthesis** | **First Author** | | | | | |
| --- | --- | --- | --- | --- | --- | --- |
|  | ***Cantey (15)*** | ***Hamdy (16)*** | ***Kilpatrick (14)*** | ***Cater (17)*** | ***Diorio (18)*** | ***Quinn (13)*** |
| **Value of AMS** In neonatal units, the involvement of AMS was not considered to be valuable by clinicians, while AS Collaboration perceived as important by QUINN in addressing NICU usage rates and practice variation.  Collaboration during rounds was considered disruptive and the presence of a dedicated pharmacist in the unit was deemed adequate.   AMS might not be relevant in all paediatric settings like oncology units. Additionally, acutely sick status of patients in secondary care, and difficulty in changing the route of administration made it difficult to implement AMS in paediatric wards. | Not valuable in neonatal units, rounds with AMS team were considered disruptive and presence of a dedicated pharmacist was enough. Antibiotic use data on dashboard was limited to higher antibiotics. |  |  | Difficult to implement AMS as pts are acutely sick.  Not applicable in paediatrics where PO administration may not be doable | AMS not relevant in paediatric oncology units Resistance did not have an impact on individual patient decision making about inf prophylaxis | ASC was perceived as important in NICUs usage rates and practice variation |
| **Structural barriers**  like affiliation of the NICU influenced the AMS coverage. Resource allocation, especially time was a barrier in getting engagement and in implementing unit change. | Separate facility: Our ASP doesn’t include the NICU Time-consuming | Weekend and night shift inertia: Institutional/structural barrier |  |  |  | Barriers experienced by participants related to allocation of time, human resources, and finances; all are important considerations when engaging in QI work. All participants expressed that time was a barrier whether it was in getting consistent engagement throughout the collaborative or trying to implement unit changes. -Technological resources were viewed as changes that were implemented in the electronic health record via documentation, antibiotic timeouts, or development of order sets. |
| **Multidisciplinary collaboration** The nursing team preferred to follow the doctors and physicians’ guidance over that of the AMS team's. They felt reluctant to defer, question and provide input to treatment plans due to reasons such as prescriber pushback, lack of involvement by consulting services, and duplication of work between the clinical team members. Unit culture and multidisciplinary collaboration were drivers in facilitating change. | Conflict with consultants: Preference to ask the consultants and doctors for help over AMS team. | Nurses reluctant to defer a rx plan, If there's a pharmacist, the nurse didn’t feel the need to provide input Nurses feel unable to provide input to resident physicians  Consulting services may not loop in the nurse Limited role in prescribing and/or use of antibiotics (SICU) Inconsistent inclusion on rounds Central role in communication with the team, Communication with other nurses |  | Prescriber pushback: IV to PO switch, because the switch is considered in the context of discharge and prescribers consider PO less effective  Duplication of work between doctors, pharmacists and nurses |  | Unit culture and multidisciplinary collaboration were drivers in facilitating change as perceived by positive support and collaboration among nurses, physicians, and pharmacists. |
| Neonatologists were resistant to input from the AMS team. The recommendations came from adult AMS providers who lacked paediatric expertise. | No paediatric expertise Recommendations from adult providers Specialized population, neonatologists resistant to input from AMS team |  |  |  |  |  |
| Communication barriers between the nursing and AMS teams resulted in unawareness around existing AMS hospital policies.  Relations between the external facilitators and the NICU affected the effectiveness. Supporting the familiarity among the team members | Communication barriers between nursing team and ASP team |  | Unawareness around existing AMS hospital policies |  |  | Collaboration with pharmacists either with daily rounding or creating antibiotic timeouts.  EF and team assignment and scheduling: Not knowing each other prior to team assignment created a barrier and may have limited the effectiveness of facilitation.  Relation developed between the EF and the NICU affected the effectiveness of the role. |
| **Nurses' role in AMS** The role of nurses in AMS was seen as valuable, but lacking recognition. Some strategies were deemed outside the scope of nurses' practice. |  | Nurses have an essential role in AMS across paediatrics and neonates. Nurses' role included administering medications safely: Minimizing errors, ensuring timeliness, adhering to protocol | Lack of recognition of the role of nurses within AMS | Nurses thought they should play a major role in AMS. Some strategies outside the nurses' scope of practice |  |  |
| **Pt/ carer Advocacy and education** Nurses believed their role was to advocate for the patients if they were uncomfortable with the physician's treatment plans, some participants felt that they weren't very good at questioning these plans. |  | Nurses serve as advocates of patients by querying the physician's rationale when they were uncomfortable with the plan, suggest alternatives or escalate concerns. Advocating for patient, Questioning the plan, Ensuring the best route of administration | Not very good at questioning why a child is on antibiotics, Patient advocacy: providing individualized care,  facilitating a positive patient experience, or advocating for parents who care for children with AD |  |  |  |
| Paediatric nurses believed that they had an important role in educating caregivers about the AMS strategies, the rationale around them, and providing tailored guidance. |  | Educating caregivers, Explaining the rationale for and risks of antibiotics, Tailoring education for families, Reviewing discharge instructions | Our role is education: Educating family members about these strategies [they need to be] getting that understanding of why to do it, when to do it, and how to do it |  |  |  |
| **Knowledge gap**  All nurse participants expressed a gap in knowledge and a need for formal training including terminology and management strategies about AMS. Lack of knowledge about antibiotic dosages and strength was listed as another barrier to nurse involvement in AMS. Participants in the EF study also expressed a desire to have a community of learning within the collaborative, |  | Desire for formal education, Nurses educating themselves through informal learning. | Lack of the terminology knowledge Lack of formal education about AMS Education surrounding management strategies for AMS | Lack of ongoing formal education around sterile techniques as part of AMS strategy, antibiotic dosage strengths |  | Participants expressed a desire to have a community of learning within the collaborative that fostered an environment of shared learning. |
| **Perceived low consumption** The hospital AMS focus on adults, with perceived low consumption in paediatric wards and NICUs. **Perceived narrow spectrum consumption** as NICUs did not use higher antibiotics, which were a focus of the AMS. | Perceived low consumption Our ASP focuses on the adults; there's just not that much use in paediatrics in general and especially the nurseries Perceived narrow-spectrum consumption We audit the nurseries, but 9 times out of 10 it's just 48 hours of ampicillin and gentamicin, so honestly it's not a focus |  |  |  |  |  |
| **Parental influence & Pushback:** Nurses in paediatric wards faced some pushback from parents when implementing certain strategies, encouraging non-invasive methods to obtain culture samples where invasive methods were indicated. |  |  |  | Pushback from patients’ families (encouraging non-invasive methods to obtain cultures where invasive methods are indicated) |  |  |
| Parents of children in oncology units expressed some concern over development of antimicrobial resistance. |  |  |  |  | Parents concern about the development of resistance |  |
| **Champion representation** was brought up in both papers studying the NICU setting | Champion representation Reduces resistance to AMS recommendations more buy-in since [a champion] began representing us at the monthly meeting. They have our perspective, and we were able to get some of the useless [stuff] off the report and look at the antibiotics we actually use |  |  |  |  | Physician and nurse champions from the three NICUs were highly engaged; however, the engagement was perceived not to move beyond local leadership |
| Individual capacity to change influenced the facilitation of AMS. |  |  |  |  |  | Individual capacity to change affected the facilitation of antibiotic stewardship |
| Neonatal teams had an increased buy-in when presented with evidence behind the patient-oriented benefit of AMS. Paediatric nurses indicated that their role in AMS was to administer medications safely, minimize errors, ensure timeliness and adhere to protocol. | Patient-level outcomes: More buy-in on reference to evidence in practice without adding to the busy work |  |  |  |  |  |
| **Monitoring and feedback** Monitoring, evaluation, and feedback was perceived as important in NICU settings. Lack of accountability to follow guidelines about proper techniques in some AMS interventions in paediatric wards was seen as a barrier. |  |  |  | Lack of accountability regarding proper techniques |  | the importance of evaluation and feedback to implementation team members, staff, families, or executive leadership. |

Supplementary Material: Example search strategy

Ovid MEDLINE(R) <1946 to present>

1	exp Antimicrobial Stewardship/

2	exp Anti-Infective Agents/ or exp Coinfection/ or exp Antimicrobial Stewardship/ or exp Drug Resistance, Bacterial/ or exp Bacterial Infections/ or Anti-Bacterial Agents/ or exp Drug Utilization/

3	((reduc$ or decreas$ or review$ or improv$) adj3 (prescrib$ or prescrip$)).tw,kf.

4	1 or 2 or 3

5	exp Infant/ or exp Child/ or exp Child, Preschool/ or paediatric*.mp. or exp Pediatrics/ or exp Adolescent/

6	Infant, Newborn/

7	neonate*.tw,kf.

8	5 or 6 or 7

9	exp Parents/

10	(parent$ or mother$ or father$ or mum$ or mom$ or dad$ or carer$ or caregiver$).tw,kf.

11	p*ediatrician.tw,kf.

12	(doctor$ or Dr$ or GP$ or clinician$ or (health adj2 professional$) or practioner$).tw,kf.

13	exp Physicians/ or exp "Attitude of Health Personnel"/

14	exp Intensive Care Units, Pediatric/ or exp Critical Care/

15	exp Inpatients/

16	exp Hospitals/

17	exp Secondary Care/

18	exp Intensive Care Units, Pediatric/

19	(P*ediatric Intensive Care or PICU).tw,kf.

20	Intensive Care Units, Neonatal/

21	14 or 15 or 16 or 17 or 18 or 19 or 20

22	exp Qualitative Research/

23	exp Interview, Psychological/ or exp Interview/

24	exp Focus Groups/

25	(narration or narrative$).tw,kf.

26	(attitude$ or view$ or belief$ or viewpoint$ or perception$ or standpoint$).tw,kf.

27	(Nudist or Nvivo or Atlas ti).tw,kf.

28	("behavioural research" or "behavioral research").tw,kf.

29	("qualitative research" or "qualitative study" or "qualitative method" or "qualitative methodology" or "qualitative design" or "qualitative methods").tw,kf.

30	("grounded theory" or "action research" or "content analysis" or "thematic analysis").tw,kf.

31	psycholog$.tw,kf.

32	ethnopsycholog$.tw,kf.

33	survey$.tw,kf.

34	("audiotape recording" or "tape recording" or taperecording* or "tape recorded").tw,kf.

35	"qualitative data".tw,kf.

36	anthropolog$.tw,kf.

37	22 or 23 or 24 or 25 or 26 or 27 or 28 or 29 or 30 or 31 or 32 or 33 or 34 or 35 or 36

38	exp Nurses/

39	(nurs* or nurs* staff).tw,kf.

40	9 or 10 or 11 or 12 or 13 or 38 or 39

41	4 and 8 and 21 and 37 and 40
