## Supplementary figures and images for "Views and experiences of Antimicrobial Stewardship interventions in paediatric secondary care settings: A Qualitative Evidence Synthesis"

### PRISMA flowchart

Figure 1: PRISMA flow chart of inclusion, literature search and study descriptions.

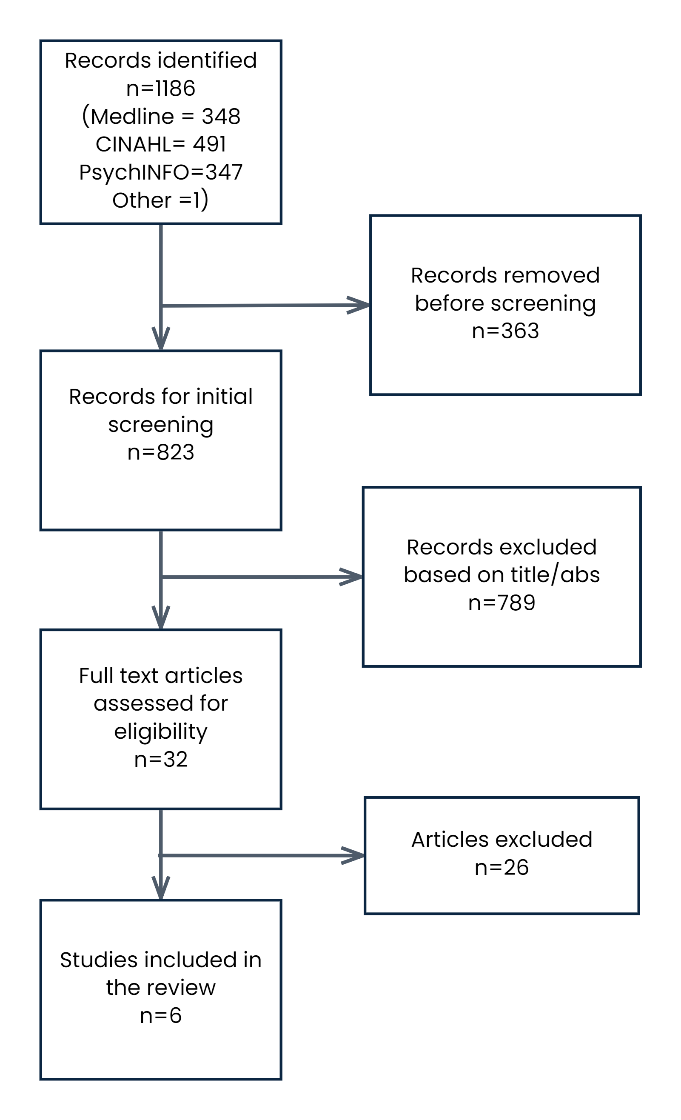
